# Less severe course of COVID-19 is associated with elevated levels of antibodies against seasonal human coronaviruses OC43 and HKU1 (HCoV OC43, HCoV HKU1)

**DOI:** 10.1101/2020.10.12.20211599

**Authors:** Martin Dugas, Tanja Grote-Westrick, Richard Vollenberg, Eva Lorentzen, Tobias Brix, Hartmut Schmidt, Phil-Robin Tepasse, Joachim Kühn

**Affiliations:** Institute of Medical Informatics, University of Münster, Germany; Institute of Virology, Department of Clinical Virology, University Hospital Münster, Germany; Medizinische Klinik B (Gastroenterologie, Hepatologie, Endokrinologie, Klinische Infektiologie), University Hospital Münster, Germany

## Abstract

The clinical course of COVID-19 is very heterogeneous: Most infected individuals can be managed in an outpatient setting, but a substantial proportion of patients requires intensive care, resulting in a high rate of fatalities. Recently, an association between contact to small children and mild course of COVID-19 was reported. We performed an observational study to assess the impact of previous infections with seasonal coronaviruses on COVID-19 severity. 60 patients with confirmed COVID-19 infections were included (age 30 - 82 years; 52 males, 8 females): 19 inpatients with critical disease, 16 inpatients with severe or moderate disease and 25 outpatients (age and gender matched to inpatients). Patients with critical disease had significantly lower levels of HCoV OC43- (p=0.016) and HCoV HKU1-specific (p=0.023) antibodies at the first encounter compared to other COVID-19 patients. Our results indicate that previous infections with seasonal coronaviruses might protect against a severe course of disease. This finding should be validated in other settings and could contribute to identify persons at risk before an infection.

## Introduction

At present, approximately 10 to 20 percent of COVID-19 patients need medical treatment in hospitals and about 5% need long-term treatment on intensive care units (ICU). In contrast, the majority of COVID-19 patients can be managed in an outpatient setting. Known important risk factors are age, male gender, high body mass index and pre-existing chronic conditions [1]. However, also young and seemingly healthy individuals are at risk to die from COVID-19 infections. At present, this heterogeneity of the disease course is not well understood. Partial immunity against SARS-CoV-2 might contribute to this phenomenon, as recently discussed in reports about cross-reactivity against SARS-CoV-2: Grifoni [2] and Le Bert [3] described T cell responses to SARS-CoV-2 in unexposed human individuals. In a recent survey, patients with mild course of COVID-19 reported frequent contact to small children [4]; therefore, exposure to childhood-related infections might modify the disease severity of COVID-19. This corresponds to the low incidence of severe COVID-19 infections in small children [5]. Seasonal coronaviruses mainly cause mild respiratory tract infections and are frequently found in children. Like SARS-CoV-2, those viruses belong to the subfamily of Orthocoronavirinae. Hence, the objective of this work is to assess if previous infections with a seasonal coronavirus – as indicated by antibody levels – modify the disease course of COVID-19.

## Material and Methods

### Patient cohort

Serum samples from 60 Patients with COVID-19 infections confirmed by RT-qPCR were analyzed in the context of the Coronaplasma Project (local ethics committee approval: AZ 2020-220-f-S) and COVID-19 biomarker study (local ethics committee approval: AZ 2020-210-f-S) at the University Hospital of Münster, Germany. Median age of patients was 58 years (range 30 - 82 years). 52 males and 8 females were included. Outpatients were manually selected to match age and gender of inpatients. After informed consent, serum was collected at the first patient contact. Median age was 58 years for outpatients (22 male, 3 female), 58 years for inpatients with critical disease (ICU group: 17 male, 2 female) and 55 years for inpatients with severe or moderate disease (non-ICU group: 13 male, 3 female), respectively. Critical disease was defined by invasive ventilation or ECMO therapy; severe disease by oxygen insufflation; moderate disease by hospitalization for other reasons without oxygen treatment. Median length of stay (LoS) for all inpatients was 10 days (range 2 - 55); 3 fatalities occurred.

### Antibody measurement

Antibodies against seasonal coronaviruses and SARS-CoV-2 were measured with the immunostrip assay recomLine SARS-CoV-2 IgG from Mikrogen GmbH, Neuried, Germany. Regarding seasonal coronaviruses, this assay measures IgG antibodies directed against the nucleocapsid protein (NP) of HCoV 229E, NL63, OC43 and HKU1. With respect to SARS-CoV-2, NP-specific and spike protein (S)-specific antibodies directed against the S1 subunit and the receptor binding domain (RBD) were determined. Measurements were performed at the Institute of Virology/Department of Clinical Virology of the University Hospital Münster according to the guidelines of the manufacturer. To test precision and reliability, a dilution series and repetitive antibody measurements were performed. Negative and positive controls were analyzed.

### Data processing and analysis

Antibody levels were visually determined according to the guidelines of the manufacturer as ordinal values using the cutoff band of immunstrips as internal reference. Results of individual coronavirus-specific bands were rated on an ordinal scale as non-detectable (−), below cutoff (+/−), with cutoff intensity (+), above cutoff (++), and very strong intensity (+++). In addition, relative antibody levels were quantitatively determined with ImageJ (version 153, 64bit-Version for windows) [6] using the signal intensity of the cutoff band of individual immunstrips as internal reference. Results were expressed as ratio signal intensity HCoV-specific band to signal intensity cutoff band. Standardized photographs from immunostrip assays were used for this analysis. Demographical data, type of treatment and length of stay were extracted by the Medical Data Integration Center (MeDIC) from the hospital information system of the University Hospital Münster (ORBIS from Dedalus Healthcare Group). Descriptive statistics and statistical tests were performed with R (version 3.6.1). Ordinal and numerical values were analyzed with Wilcoxon tests. Association of numerical values was assessed with Spearman correlation. A two-sided p-value of 0.05 was considered significant.

## Results

According to the visual determination of band intensities, levels of HCoV OC43- and HKU1-specific IgG antibodies were significantly lower for COVID-19 inpatients with critical disease compared to all other patients (p=0.016 for OC43; p=0.023 for HKU1; Wilcoxon test with ordinal measurement data). Figure 1 presents ordinal IgG antibody levels against HCoV for COVID-19 patients with critical disease compared to other COVID-19 patients. This finding was confirmed by densitometric determination of relative antibody levels (Figure 2).

**Figure 1:**
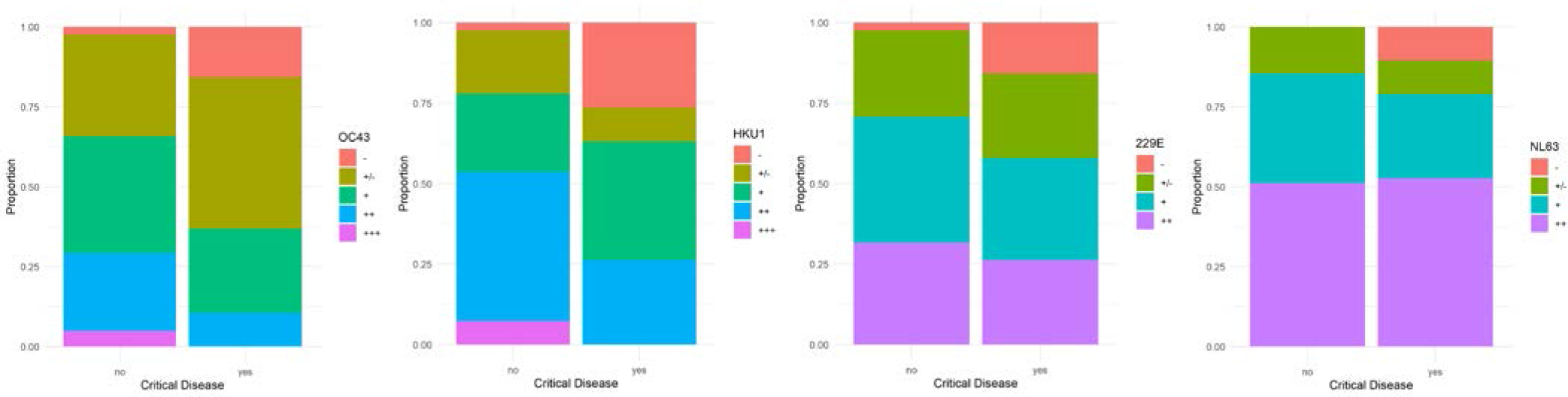
Proportion of ordinal HCoV antibody levels from COVID-19 patients with and without critical disease. a) OC43 (p=0.016) b) HKU1 (p=0.023) c) 229E (p=0.30) d) NL63 (p=0.82). COVID-19 patients with critical disease present low antibody levels more frequently than patients without critical disease. This difference is significant for OC43 and HKU1.

**Figure 2:**
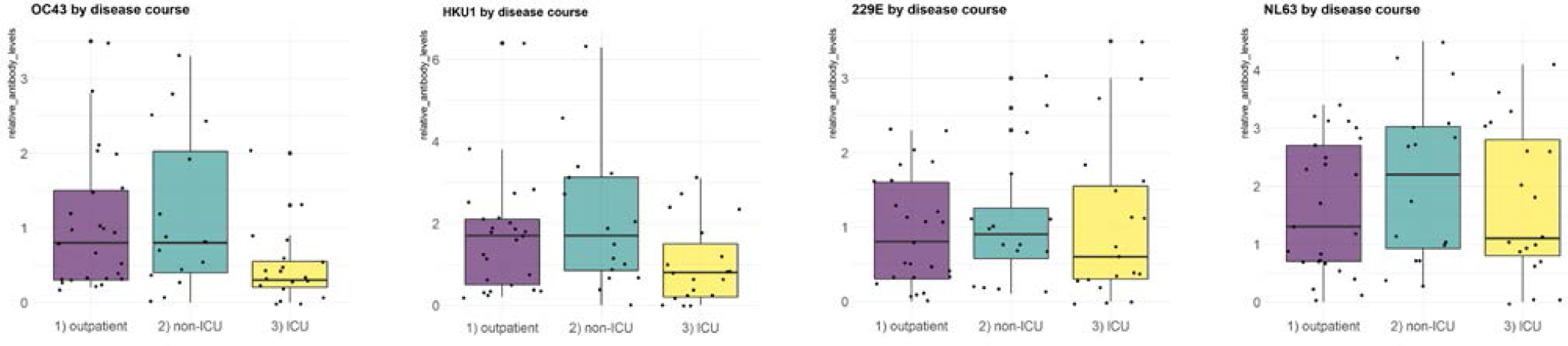
Relative HCoV antibody levels of OC43 (a), HKU1 (b), 229E (c), NL63 (d). COVID-19 patients with critical disease (ICU group) present lower values than other groups. This difference is higher for OC43 and HKU1 than for 229E and NL63.

To further assess potential clinical implications of antibodies against endemic coronaviruses for COVID-19 patients, correlation of LoS and antibody levels against HCoVs OC43 as well as HKU1, respectively, was analyzed (Figure 3). Long hospitalization periods were predominantly seen in patients with low antibody levels, but this correlation was not significant.

**Figure 3:**
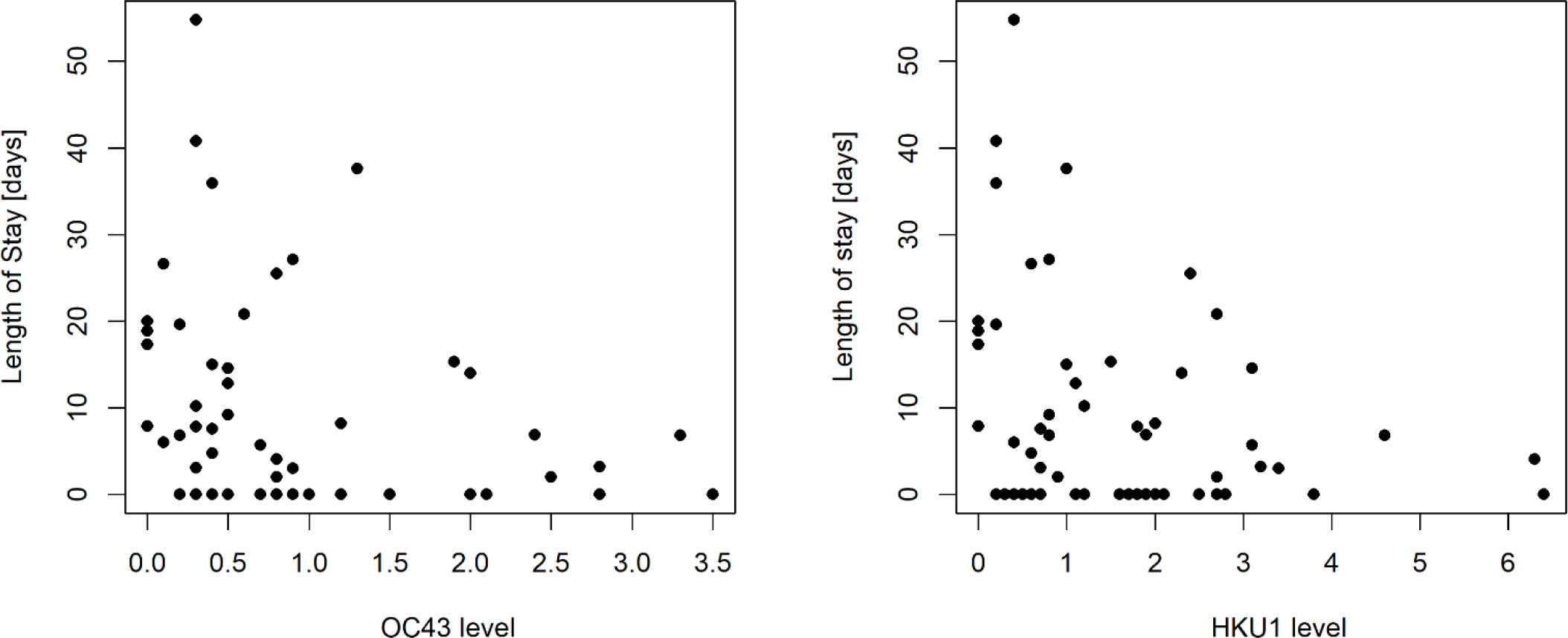
a) Correlation of length of stay and OC43 antibody levels. A trend is visible, but not significant (p=0.068). b) Similar result for correlation of LoS and HKU1 antibody levels. A trend is visible, but not significant (p=0.083). Higher antibody levels are correlated with reduced duration of hospitalization.

Figure 4 presents results from SARS-CoV-2 IgG antibody measurements at first encounter. In general, patients with critical disease had higher SARS-CoV-2 IgG antibody levels compared to moderate/severe inpatients and outpatients.

**Figure 4:**
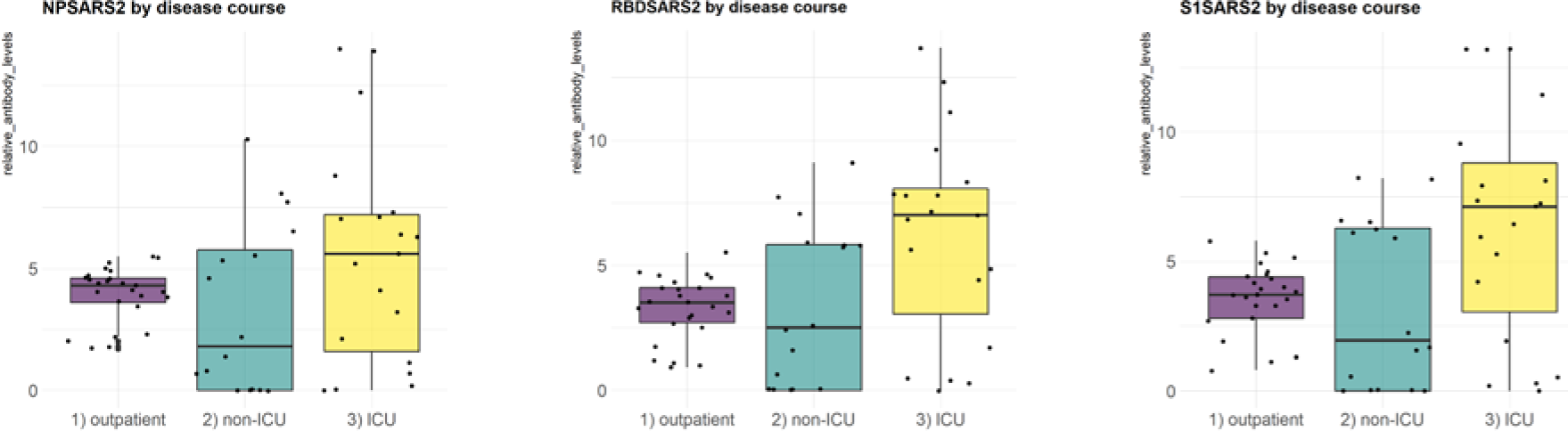
Boxplots of SARS-CoV-2 relative antibody levels for outpatients, patients with severe/moderate disease (non-ICU group) and critical disease (ICU group). Median antibody levels are higher for patients with critical disease. a) NP SARS-2 b) RBD SARS2 c) S1 SARS2

## Discussion

Recently, several groups have reported immunological cross reactivity against SARS-CoV-2 in unexposed individuals [2, 3]. From a public health perspective, the relatively high proportion of COVID-19 patients with critical disease poses the key problem of this pandemic: overload of the healthcare system. If a previous infection with a known pathogen modified the course of COVID-19, i.e. decreased the proportion of intensive care patients, this could become a component to overcome the pandemic: Persons at risk for a severe course of disease could be identified *before* a SARS-CoV-2 infection and appropriate protective measures could be taken. Of note, this might also be relevant for vaccination strategies.

This observational study assessed a potential relationship between previous infections with seasonal coronaviruses – measured as antibody levels – and the severity of COVID-19 disease. It was shown that elevated antibody levels for HCoVs OC43 and HKU1 were associated with less need for intensive care therapy. In addition, a clear trend towards a reduced length of hospital stay was observed. One might argue that higher levels of antibodies against seasonal coronaviruses are merely a surrogate marker for a more active immune system. However, HCoVs OC43 and HKU1 are betacoronaviruses and therefore closer related to SARS-CoV-2 than HCoVs 229E or NL63. This is in line with our data that previous exposure to HCoVs OC43 and HKU1 has a stronger association with severity of COVID-19 than HCoV 229E or NL63 infections in the past. A possible explanation might be that previous exposure to seasonal betacoronaviruses facilitates immune response to SARS CoV-2. Further research is needed to assess the molecular mechanism behind our findings. Of note, cross-reactivity between HCoV OC43 and SARS-CoV was already described in 2006 [7]. Kissler [8] developed a simulation model for transmission dynamics of SARS-CoV-2 and reports that even mild cross-immunity from HCoV OC43 and HCoV HKU1 could potentially have a relevant effect on SARS-CoV-2 transmission.

Regarding SARS-CoV-2 antibodies, an inverse pattern was detected: inpatients with critical disease demonstrated higher median antibody levels for NP, RBD and S1 of SARS CoV-2 compared to other patients. Similar results were reported recently [9]. Hence, there is no evidence for a general bias regarding antibody levels and the ability to mount a humoral immune response between the three groups (outpatient, non-ICU, ICU) in our cohort.

This study has important limitations: It is a retrospective single-site study with a limited number of cases and association is not causation. However, it is remarkable that the effect of HCoV OC43 and HKU1-specific antibody levels reached statistical significance regarding the need for intensive care therapy with only 60 patients.

Therefore, these findings should be validated in other sites with larger patient collectives. In a prospective setting (e.g. for risk groups with contacts to many persons like employees of hospitals or supermarkets) it should be tested if the absence of HCoV OC43 and HKU1-specific antibody levels can identify persons at risk for a severe course of COVID-19. Identification of vulnerable individuals is a key priority in the current stage of the pandemic to guide protective measures and to design vaccination strategies.

## Conclusion

Elevated levels of pre-existing antibodies against seasonal coronaviruses, specifically HCoV OC43 and HKU1, are associated with less severe course of COVID-19. Further studies should validate this finding and explore the potential to identify persons at risk for severe disease course before a SARS-CoV-2 infection.

## Data Availability

Due to data protection regulations, personal identifiable data cannot be published.

## Acknowledgement

Supported by a grant from BMBF (HiGHmed 01ZZ1802V, Use Case Infection Control).

## Data availability statement

Due to data protection regulations, personal identifiable data cannot be published.

## Code availability statement

R code is available on request from the authors.

## Notes

### Competing Interest Statement

The authors have declared no competing interest.

### Author Declarations

local ethics committee approval: AZ 2020-220-f-S and AZ 2020-210-f-S

